# Ranking videolaryngoscopes by orotracheal intubation performance: protocol of a systematic review and network meta-analysis of clinical trials at patient level

**DOI:** 10.1101/2021.01.18.21250062

**Authors:** Clístenes Crístian de Carvalho, Danielle Melo da Silva, Victor Macedo Lemos, Thiago Gadelha Batista dos Santos, Ikaro Cavalcante Agra, Gustavo Miná Pinto, Isabella Beserra Ramos, Yuri Soares da Cunha Costa, Jayme Marques dos Santos Neto

**Affiliations:** Department of Post-graduation, Instituto de Medicina Integral Professor Fernando Figueira, Recife, Brazil; department of surgery, Universidade Federal de Campina Grande, Campina Grande, Brazil; Support and Therapeutic Diagnosis Division, Anesthesiology and post-anesthetic care unit, Hospital das Clínicas da Universidade Federal de Pernambuco, Recife, Brazil; Centro de Ciências Biológicas e da Saúde, Universidade Federal de Campina Grande, Campina Grande, Brazil

## Abstract

**Background:** Videolaryngoscopes (VLs) are regarded to improve glottic visualization as compared to Macintosh laryngoscope (ML). However, we currently do not know which one would be the best choice. We then designed this systematic review and network meta-analysis to rank the different VLs as compared to ML.

**Methods:** We will conduct a search in PubMed, LILACS, Scielo, Embase, Web of Science, and Cochrane Central Register of Controlled Trials (CENTRAL; 2020, Issue 6) on 11/01/2021. We will include randomized clinical trials fully reported with patients aged ≥ 16 years, comparing VLs with ML for failed intubation with the device, failed first intubation attempts, number of intubation attempts, time for intubation, difficulty of intubation, and improved visualization of the larynx. Pooled effects will be estimated by both fixed and random-effects models and presented according to qualitative and quantitative heterogeneity assessment. Sensitivity analyses will be performed as well as a priori subgroup, meta-regression and multiple meta-regression analyses. Additionally, network meta-analyses will be applied to rank the different VLs as compared to ML. We will also assess the risk of selective publication by funnel plot asymmetry.

**Discussion:** This systematic review and network meta-analysis aim at helping health services and clinicians involved in airway manipulation choose the best VLs for orotracheal intubation.

**Systematic review registration:** The current protocol was submitted to PROSPERO on 07/01/2021.

## INTRODUCTION

Intubation hardships were shown to be the most frequent primary airway problems during anaesthesia – 39%^1^. Even so, despite the importance of predicting difficult intubations in order to better manage these airways, we are currently not able to accurately predict such scenarios – much because of the unreliable predictive values of available diagnostic tests^2-4^. As a consequence, anaesthesiologists fail to anticipate around 93% of difficult intubations, placing these patients at higher risk of life-threatening complications^5^.

As an alternative to better approach patients with difficult direct laryngoscopy, we may benefit from the use of videolaryngoscopes (VLs). Studies have suggested the use of these devices to improve the view of the larynx during laryngoscopy, what might enhance chances of successful tracheal intubation for patients with difficult direct laryngoscopy^6-8^. A meta-analysis that compared VLs – indistinctly – with direct laryngoscope with Macintosh blade (ML) showed the former to outperform the latter for failed intubation, airway trauma, hoarseness, difficulty of intubation, and improved larynx visualization^9^. However, authors did not compare VLs directly, although they have found evidence of differential performance between different designs. This way, even though available evidence suggests VLs to outperform ML for tracheal intubation, we currently do not know whether a particular device might be chosen over the others.

Therefore, we designed this systematic review and network meta-analysis in an attempt to rank VLs for orotracheal intubation performance as compared to ML. For this purpose, we will assess the influence of each laryngoscope on failed intubation with the device, failed first intubation attempts, number of intubation attempts, time for intubation, difficulty of intubation, and improved visualization of the larynx.

## METHODS

### Protocol and registration

The current protocol was designed according to recommended standards and reported as per the Preferred Reporting Items for Systematic Reviews and Meta-analysis Protocols (PRISMA-P) guidelines^10-12^. This review protocol was also submitted to PROSPERO registration on 07/01/2021.

### Eligibility criteria

Inclusion criteria will be as follows: 1) randomized clinical trials fully reported; 2) patients aged ≥ 16 years – not manikins; 3) data available on failed intubation with the device, failed first intubation attempts, number of intubation attempts, time for intubation, difficulty of intubation, and improved visualization of the larynx; 4) comparison between VLs or between these and ML. Exclusion criteria will be as follows: 1) studies published in language other than English, Spanish or Portuguese; 2) impossibility of abstracting relevant data on outcomes at patient level; 3) differences in the intubation technique, including drugs used, between intervention groups other than the laryngoscopes.

### Information sources

We will conduct a computerized search through PubMed, LILACS, Scielo, Embase, Web of Science, and Cochrane Central Register of Controlled Trials (CENTRAL) on 11/01/2021. We will also search the reference lists of included studies.

### Search strategy

The following searching strategy line will be applied to databases with no limitations: *(laryngoscopes[MeSH] OR laryngoscop* OR videolaryngoscop* OR GlideScope OR Pentax OR C-MAC OR blade OR McGrath OR X-lite OR Airtraq OR Trueview OR CEL-100 OR “King vision” OR Bullard OR Venner OR vividtrac OR “copilot VL” OR “ue?scope”) AND (“Airway management”[MeSH] OR “Airway management” OR intubation* OR difficult* OR visualization OR view)*

### Study records

Retrieved references will be taken to EPPI Reviewer Web (Beta)^13^ for screening steps – “title and abstract” then “full text”. Eligibility criteria will be applied to select the studies to be included. Four doubles of reviewers will perform in duplicate and independently all steps from screening of title and abstract, through screening of full text and risk of bias assessment to data extraction. The results will be compared and disagreements solved by discussion and consensus amongst correspondent researchers and CC. In case of no consensus, CC will act as final judge. In case of missing data, corresponding authors will be reached. If relevant data is missing or data are conflicting and corresponding author do not reply our contact after three attempts, then the article will be excluded. Data will be recorded in an excel spreadsheet. The data-extraction form will first be tested in 5 included studies and then refined if necessary.

### Data items

We will collect data on first authors name, publication year, study design, characteristics of population, mean age, mean BMI, mean weight, mean height, sex frequencies, ASA physical status, setting, country, sampling, nature of procedure (elective vs urgent), intubation technique (regular vs rapid sequence induction vs awake), manipulator experience, number of participants randomised and analysed, number of participants in each arm, type of laryngoscope, inducer used and dose, opioid used and dose, neuromuscular blocking agent used and dose. Also, for continuous outcomes, mean and standard deviation in each arm and mean difference and standard error between groups. For categorical outcomes, number of events and number of patients in each arm along with relative risk and standard error between groups.

### Outcomes with prioritization

We will primarily investigate the risk of failed intubation with the devices – which authors consider the most important outcome when defining intubation performance, mainly for difficult intubations. To getting additional information about how different laryngoscopes perform during orotracheal intubation attempts, we will also evaluate their effects over failed first intubation attempts, number of intubation attempts, time for intubation, difficulty of intubation, and improved visualization of the larynx.

### Risk of bias in individual studies

We’ll apply the RoB 2 tool to assessing the risk of bias of the individual studies for each outcome^14^. Five domains are assessed through this tool: randomization process; deviation from intended interventions; missing outcome data; measurement of the outcome; and selection of reported results. We will also use risk of bias judgments for sensitivity analysis.

### Data synthesis

Data will be summarized if at least two different sources are available. Analyses will be conducted in Review Manager^15^ (RevMan, London, UK, v5.3.5) and R software tools^16^ (R Foundation for Statistical Computing, Vienna, Austria), as appropriate. Per-protocol data on patient level will be extracted or calculated from studies and used for summarizations. Effect sizes, SE, and 95% CI will be estimated for each study from the recorded data. Forest plots of relative risk or mean difference will be constructed for every outcome. Pooled estimates will be calculated by both fixed-effects (Mantel-Haenszel or inverse variance method, where appropriate) and random-effects (Sidik-Jonkman method with Hartung-Knapp adjustment) for sensitivity analyses. Heterogeneity will be evaluated qualitatively and quantitatively by Cochran’s Q-test, ***I***^2^, and Graphic Display of Heterogeneity (GOSH). Where qualitative or quantitative heterogeneity is present, pooled estimates from random-effects models will be presented. An influence analysis by Leave-One-Out method will be performed to assess the influence of each study in the pooled effects and the between studies heterogeneity. GOSH plots will also be used to search for subclusters of different effects sizes in order to support the subgroup evaluations. Sensitivity analyses will be undertaken throughout both subgroup and meta-regression analyses. Subgroup assessments will be performed using either mixed-effects or random-effects models, where appropriate, for outcomes with 10 or more studies available. Meta-regressions of single features, one at a time, as well as multiple meta-regression with maximum likelihood (ML) estimator for *τ*^2^ will be conducted only for outcomes with 10 or more studies available per covariate. The multiple meta-regression models will be submitted to a permutation test to confirm statistical significance. Both subgroup and meta-regression analyses will be performed with a priori hypotheses – attempting to avert the catch of spurious associations – with the following features: manipulator experience, intubation technique, population characteristics, setting, nature of procedure, type of laryngoscope, inducer, opioid, and neuromuscular blocking agent used. Risk of bias judgments and decisions made throughout the statistical analysis will also be included in sensitivity analyses. Random-effects network meta-analyses among the different VLs and ML will also be performed. Network models’ consistency will be tested by net heat plots and net splitting method. Additionally, to deal with the risk of type-I and type-II errors due to repeated significance testing by subsequent meta-analyses, we will apply the trail sequential analysis for the main outcome.

### Meta-bias

We will perform assessment of selective publication by small sample bias methods for those outcomes with 10 or more studies. Funnel plots will be built and Egger’s tests performed to check for plot asymmetry. The threshold of significance will be set at p<0.100 for this method as this test has low power. Where asymmetries are present, a Duval and Tweedie’s trim-and-fill procedure will be applied to estimate bias-corrected effects.

### Confidence in cumulative evidence

To assess the quality of the evidences, we will apply the Grading of Recommendations, Assessment, Development and Evaluation (GRADE) approach. This approach takes into account factors related to design of the study, risk of bias, inconsistent results, indirectness of evidence, imprecision, publication bias, the magnitude of the effect, existence of dose-response gradient, and if all plausible confoundings would only reduce the demonstrated effect.

## DISCUSSION

This systematic review and network meta-analysis aim to evaluate whether available evidence is enough to justify the choice of some VLs over the others. Current evidence suggests VLs to outperform ML for tracheal intubation and different VLs designs to have differential performance. However, we are not yet grounded to choose a particular device or set of devices among those available. We will then attempt to rank VLs as compared to ML by a network meta-analysis in order to help health services and clinicians involved in airway manipulation choose the best VLs for orotracheal intubation.

## Data Availability

Not applicable

